# Migrant children participation during emergencies: A Scoping review of global challenges and opportunities

**DOI:** 10.1101/2024.01.09.24300971

**Authors:** Bassma Aammar, Pascale Salameh, Jose Isaias Garcia Ulerio

**Author notes:** **Corresponding author:** Bassma Aammar.

## Abstract

**Introduction:** Children who are internally displaced or migrants face severe difficulties during emergencies, including exposure to violence, exploitation, disrupted education, and mental health issues. The principle of child participation, enshrined in the United Nations Convention on the Rights of the Child (UN, 1992), recognises children as rights holders with the ability to actively engage in decision-making processes. However, the literature on child participation in emergency response and recovery efforts for these vulnerable populations remains limited.

**Objective:** The aim of this scoping review is to map the existing literature regarding child participation in emergencies. Consecutively, we want to identify opportunities and challenges related to child participation in emergency response and recovery interventions for migrant or internally displaced children (IDP).

**Methodology:** A systematic search was performed through PubMed, Scopus, Web of Sciences, and Google Scholar by including key terms related to child participation, emergencies, IDPs, and migrants. The inclusion criteria focused on papers that analyse and conclude on the opportunities and challenges within three core domains: health promotion, education integration, and social integration. Qualitative and quantitative studies published between 2003 and 2023 were included. No geographical limitations were applied to ensure a global perspective. Lastly, data extraction was carried out using the evidence synthesis software RAYYAN under the PRISMA flowchart to provide a methodical collection and presentation.

**Results:** The scoping review identified a total of 20 articles, showcasing both challenges and opportunities related to child participation in emergencies. Across the articles, health promotion was addressed in 3 articles, education integration in 14 articles, and social integration in 16 articles. The challenges encompassed issues such as limited access to healthcare, education, and social services, as well as discrimination and a lack of documentation. Opportunities included the provision of youth-centred healthcare, strengths-based approaches, intersectoral collaboration, and the empowerment of children to actively participate in decision-making processes. Challenges and opportunities were context-dependent but collectively emphasised the importance of considering the multifaceted aspects of child participation in emergency settings.

**Conclusion:** This scoping review provides insights into the landscape of child participation in emergencies, particularly focusing on health promotion, education integration, and social integration. The challenges identified underscore the barriers faced by migrant and internally displaced children in accessing essential services and their struggle for social inclusion. In contrast, the opportunities discussed shed light on promising approaches to overcome these challenges, highlighting the significance of youth-centred services, resilience-building, and intersectoral collaboration.

The findings not only contribute to the existing literature but also emphasise the importance of recognising children as active agents of change in emergency situations, aligning with the principles of the United Nations Convention on the Rights of the Child. Future research and policy initiatives should aim to further enhance child participation and integration in emergency response and recovery efforts.

## Introduction

Migrant and internally displaced children represent a substantial segment of populations affected by humanitarian crises, facing multifaceted challenges during emergencies, including physical and emotional trauma, exploitation, and disrupted education [1] [2]. The United Nations Refugee Agency reports that 41% of forcibly displaced individuals are children under 18, emphasizing the critical role of their involvement in disaster preparedness[3].

The third pillar of the United Nations Convention on the Rights of the Child emphasizes child participation, advocating for their active involvement in decision-making processes during crise[4]. However, the literature on child participation in emergency response and recovery efforts for these vulnerable populations remains limited.

This scoping review aims to map the landscape of child participation during emergencies, particularly focusing on health promotion, School integration, and social inclusion. By delineating challenges and identifying opportunities, this review seeks to inform policies and initiatives that prioritize the rights, well-being, and resilience of migrant or internally displaced children during crises.

## Objectives

The primary aim of this scoping review was to comprehensively map and synthesize existing literature on child participation in emergency response and recovery among internally displaced or migrant children. Specifically, the objectives included identifying challenges and opportunities encountered by these vulnerable populations in various emergency settings globally.

## Methodology

This study utilized a scoping review methodology to comprehensively map existing literature, focusing on migrant child participation in emergencies. Unlike systematic reviews, which narrow down specific research questions, scoping reviews aim to identify the breadth of evidence, offering a broader perspective to explore gaps, patterns, and key concepts within the literature. A structured search strategy was employed across PubMed, Scopus, and Web of Science databases from 2003 to 2023. The search aimed to identify studies focusing on child involvement in decision-making processes, empowerment, and activities relevant to emergency preparedness and response.

### Literature Search and Information Sources

The search strategy utilized Medical Subject Headings (MeSH) terms and specific keywords related to child participation, emergencies, and associated domains. The inclusion criteria encompassed studies targeting children, adolescents, and young adults in emergency contexts across various domains. Unpublished works and studies not available in English were excluded.

### Eligibility Criteria

The inclusion criteria encompassed a wide range of study designs, including qualitative, quantitative, and mixed-methods studies, relevant to the domains of health promotion, education integration, and social integration. Studies focusing on children, adolescents, and young adults in emergency situations, examining their involvement in decision-making processes, empowerment, and activities pertinent to emergency preparedness and response, were included.

### Study Selection Process

Following the database searches, identified articles were screened based on titles, followed by the removal of duplicates using Rayyan reference management software. [5] The abstracts of the remaining articles were reviewed independently by two researchers to ensure alignment with the predefined inclusion criteria. Any discrepancies were resolved through consensus, and a full-text review was conducted to finalize article selection.

### Extraction and Synthesis

The selected articles underwent data extraction using “Appendix 11.1 of the JBI Manual of Evidence Synthesis: Template Source of Evidence Details, Characteristics, and Results Extraction Instrument.” Variables such as study design, emergency type, regional context, and intervention domains (social, educational, or health) were systematically extracted. Additionally, thematic areas focusing on approaches, usage, skills, and regulatory policies were identified and analyzed to comprehend challenges and opportunities. [6]

### Quality Assessment and Analysis

The methodological rigor and quality of the included studies were evaluated using established checklists from JBI guidelines. Moreover, a level of evidence categorization ranging from V to VII was utilized to determine the reliability and robustness of the studies. This comprehensive assessment and synthesis aim to enhance transparency in the review process. (https://libguides.winona.edu/ebptoolkit/Levels-Evidence).

## Results

The systematic search identified 254 articles across databases. After removing duplicates and screening titles and abstracts, 19 papers met the eligibility criteria and were included for analysis.(Figure 1)

**FIGURE 1:**
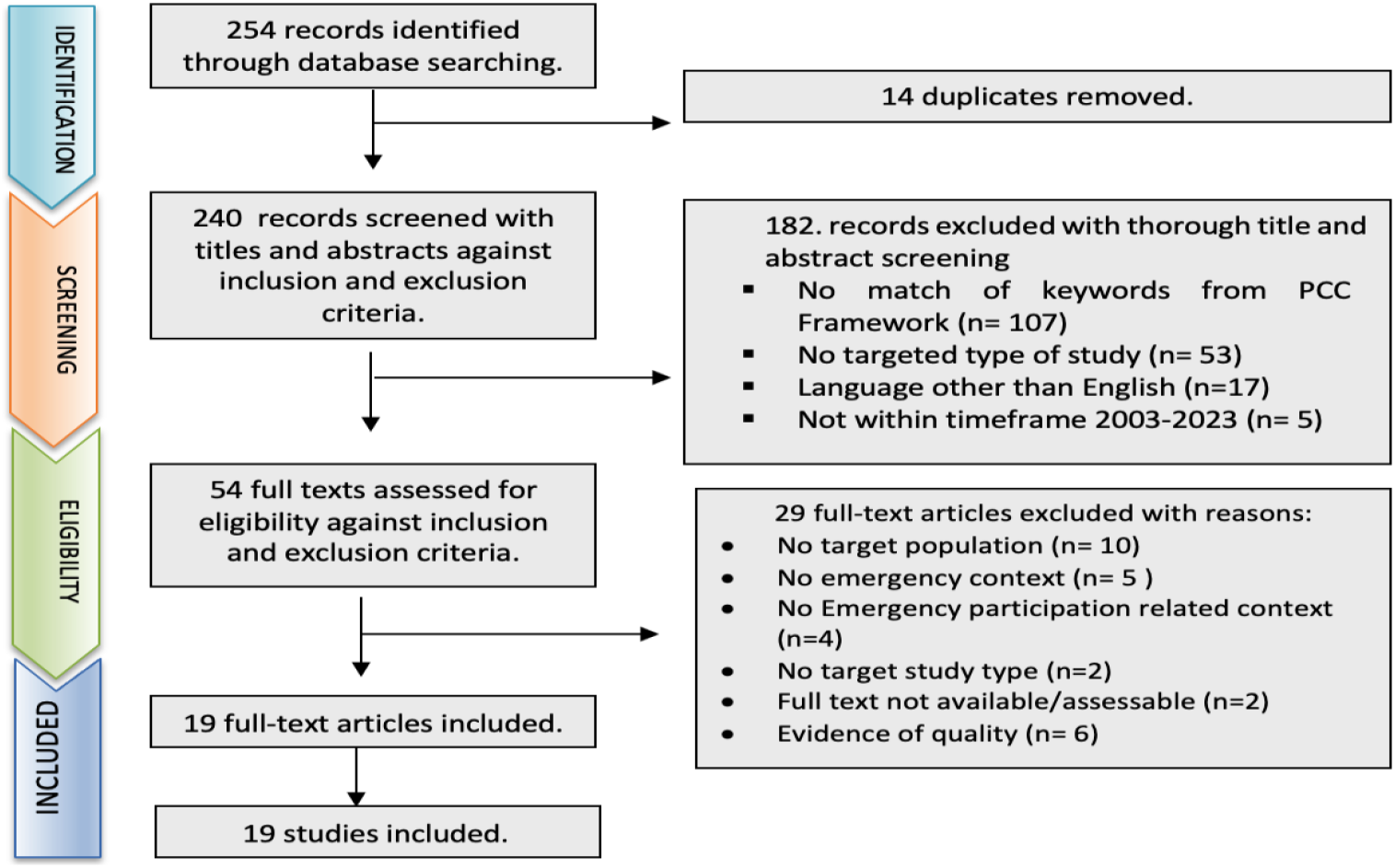
PRISMA FLOWCHART.

**FIGURE 2.**
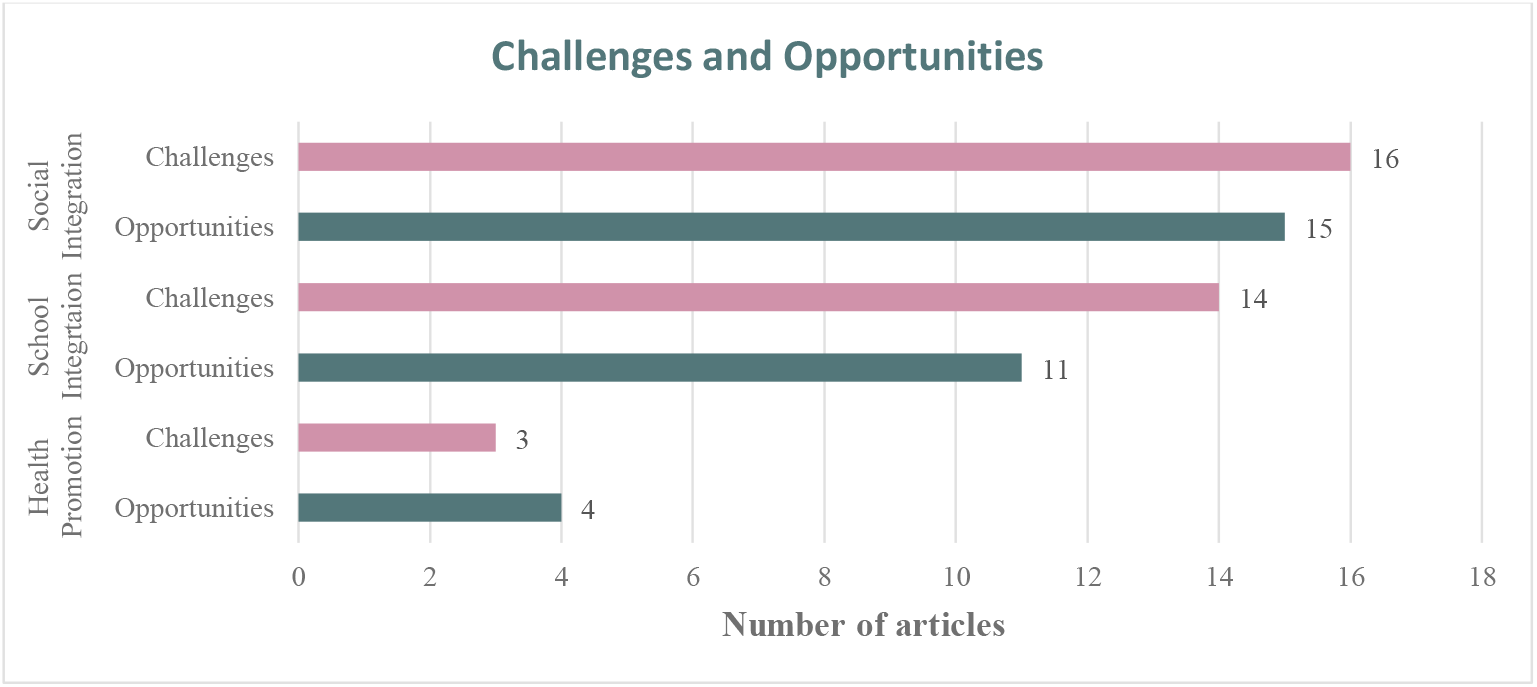
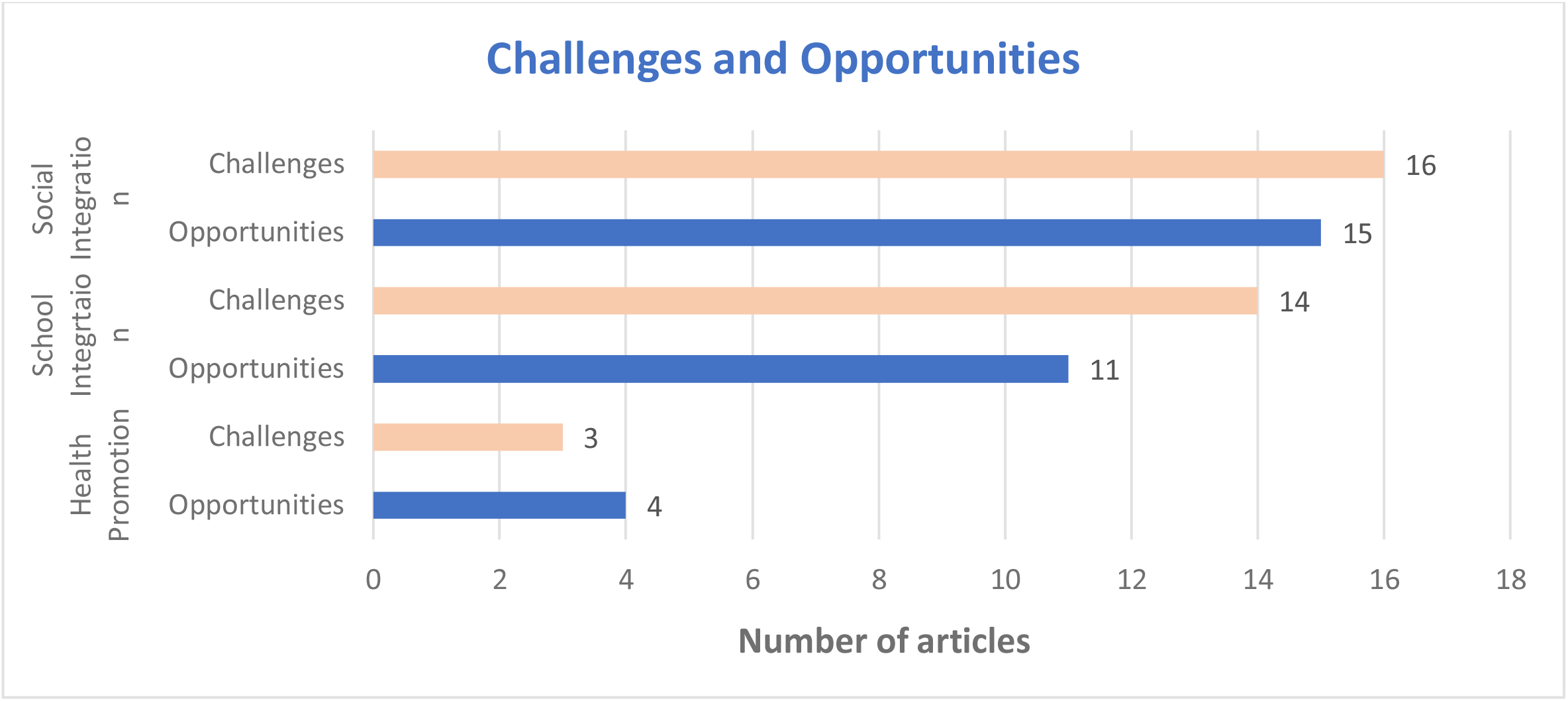
CHALLENGES AND OPPRTUNITIES.

### Study Characteristics

#### Type of Evidence

Most articles were qualitative studies (15 articles), followed by one quasi-experimental study (1 articles), a pilot study, textual evidence based on expert opinions, and textual evidence based on policies (each representing 1 article each). (Table 1)

**TABLE 1.** STUDY CHARACTERISTICS.

| Article Characteristics | Articles number |
| --- | --- |
| **Type of Evidence** |  |
| Qualitative | 15 |
| Quasi-experimental study | 1 |
| Pilot study | 1 |
| Textual Evidence: Expert opinion | 1 |
| Textual Evidence: Policy | 1 |
| **Emergency Type** |  |
| Pandemic | 6 |
| Armed Conflict | 12 |
| Forced migration | 16 |
| Natural events | 3 |
| **Region where studies conducted** |  |
| Europe | 4 |
| Asia | 2 |
| Oceania | 2 |
| Africa | 3 |
| North America | 2 |
| Middle East | 6 |

#### Emergency Type

Articles covered various emergencies, with a focus on forced migration (16 articles), armed conflict (12 articles), pandemics (6 articles), and natural events (3 articles). (Table 1)

#### Population Age and Settings

Target population included above below 15 years old and those aged 15 to 18 years. Studies primarily focused on interventions in camps (13 articles) and community host settings (10 articles). (Table 2)

**TABLE 2:** STUDY POPULATION.

| Population Age | No of Articles |
| --- | --- |
| From 5 to 15 years | 16 |
| From 15 to 18 years | 16 |
| population Setting | No of articles |
| community host | 10 |
| camps | 13 |

**TABLE 3.** EVIDENCE LEVELS.

| Evidence Levels | No of Articles |
| --- | --- |
| III | 3 |
| V | 5 |
| VI | 8 |
| VII | 1 |

#### Regions and Countries

The studies were conducted globally, with significant representation from the Middle East (6 articles), Europe (4 articles), North America, Oceania, and Asia (6 articles) and Africa (3 articles). Individual countries represented included, United Kingdom, United States, Australia, Syria, Tanzania, Sudan, South Africa, Italy, Turkey, Lebanon, El alto in Bolivia, Bangladesh, China, Germany, The West Bank, And Canada.

#### Evidence Levels

Most studies fell into Level VI (8 articles), and Level V (5 articles), followed by Level III (3 articles) and Level VII (1 article). (Table 2)

#### Challenges and Opportunities

The analysis of the included articles has revealed several critical characteristics regarding the challenges and opportunities associated with children’s participation in emergency scenarios (Figure 3) .These characteristics shed light on the prevailing trends and priorities in the field of child participation during crises.

#### Social Integratio

Migrant children faced multifaceted challenges affecting their integration, including economic disparities, language barriers, and ethnocultural discrimination. These hurdles hindered their sense of belonging and acceptance within communities. Studies by Lamb, Farini and Huang highlighted issues such as poverty, communication obstacles, and financial hardships faced by migrant children, compounded by legal complexities and spatial segregation. [7][8][9]

In contrast, various opportunities were identified. Initiatives fostering integrational resilience, trust-building among peers, community-based safe spaces, educational services, and psychological counselling emerged as pivotal tools in fostering inclusivity and social integration. [10] [11] [12] [13]

#### School Integration

Education posed significant challenges for migrant children, ranging from access constraints to language barriers and limited subject offerings within formal educational systems. Studies by Culbertson, Celik, Lamb and Prodip highlighted difficulties, including transportation challenges, early marriages, and inadequate educational resources. [14][7][15] [16]

However, opportunities exist in tailored policies for refugee students, gender-sensitive approaches, safety considerations within educational settings, and integrated mental health services. [10] [11] [17] [13]

#### Health Promotion

Migrant children encountered health-related challenges like gender-based violence, communication barriers, and limited access to healthcare services. Studies by Sommer et al., Block and Gibbs, and Huang et al. illuminated these issues, citing financial burdens, violence, and inadequate resources as hindrances. [17] [18] [9]

Conversely, opportunities centered around poverty alleviation, peer socialization, trust-building, psychological counselling, comprehensive healthcare services, and initiatives addressing gender-based violence. [7] [8] [10] [19] [13]

## Discussion

The identified challenges in social integration, education, and health promotion among migrant children during emergencies resonate with broader research findings [7][17][14]. These multifaceted hurdles, including economic disparities, language barriers, and gender-based violence, reflect the complex interplay of sociocultural factors affecting the integration of vulnerable populations.

However, the opportunities identified align with previously suggested strategies and intervention[10][11]. Initiatives promoting trust-building, community-based safe spaces, gender-sensitive approaches in education, and comprehensive healthcare services offer promising avenues to mitigate challenges faced by migrant children.

The findings of this review underscore the critical need for holistic, context-specific interventions involving policymakers, humanitarian organizations, and local communities. Tailored policies, gender-sensitive approaches, and comprehensive health and education initiatives emerge as imperative components of emergency response strategies [20] [21] [19]

### Generalizability and Limitations

While the review offers valuable insights, it is crucial to acknowledge certain limitations. The predominance of qualitative studies might limit the synthesis to certain perspectives, possibly influencing the overall findings. Additionally, while the review encompassed diverse geographical locations and emergencies, the specific contextual and cultural nuances of each region may restrict the broad applicability of the conclusions drawn.

### Implications for Public Health Practice and Future Research

Addressing the identified challenges requires a comprehensive approach involving policymakers, humanitarian organizations, and local communities. Initiatives focusing on social inclusion, tailored educational strategies, and holistic health promotion programs could significantly enhance child participation in emergencies. Future research endeavors should aim for more robust methodologies, including longitudinal studies and diverse participant demographics, to address the gaps highlighted in this scoping review.

## Conclusion

This scoping review underscores the intricate challenges faced by internally displaced or migrant children in emergency settings, encompassing social, educational, and health-related dimensions. Integrating opportunities identified, such as trust-building initiatives and tailored policies, into emergency response efforts could significantly improve the well-being and integration of these vulnerable populations. The study calls for a more nuanced approach to policymaking and research, emphasizing the need for culturally sensitive, context-specific interventions to enhance child participation in emergency response and recovery.

## Supporting information

Search Strategy

## Data Availability

All data produced in the present work are contained in the manuscript

## Acknowledgment

Pr. Pascale Salameh, Pr. Christiana Demetriou, and Dr. Jose Isaias Garcia Ulerio deserve our heartfelt gratitude for their helpful guidance, intellectual insights, and continuous support during this research endeavor. Their knowledge, encouragement, and dedication all contributed significantly to the study’s success.

---

**Annex 1:** Adjusted JBI template source of evidence.

|  |  |
| --- | --- |
| Study details |  |
| Title |  |
| Objectives |  |
| Question |  |
| Year of Publication |  |
| Authors |  |
| Inclusion / Exclusion Criteria |  |
| Language | English |
| Emergency type | Forced migration, Armed conflict, pandemics, natural events |
| Target population | Migrant / IDP or refugee Children and adolescents ( 5 to 18 years old) |
| Challenges and opportunities characteristics | Scholl integration, health promotion and social inclusion |
| Settings | Refugee camps, Host communities |
| Study outcomes / results extracted |  |
| Study outcomes |  |
| Study limitation / biases |  |
| Quality assessment |  |
| Levels of evidence |  |

**Annex 2:**
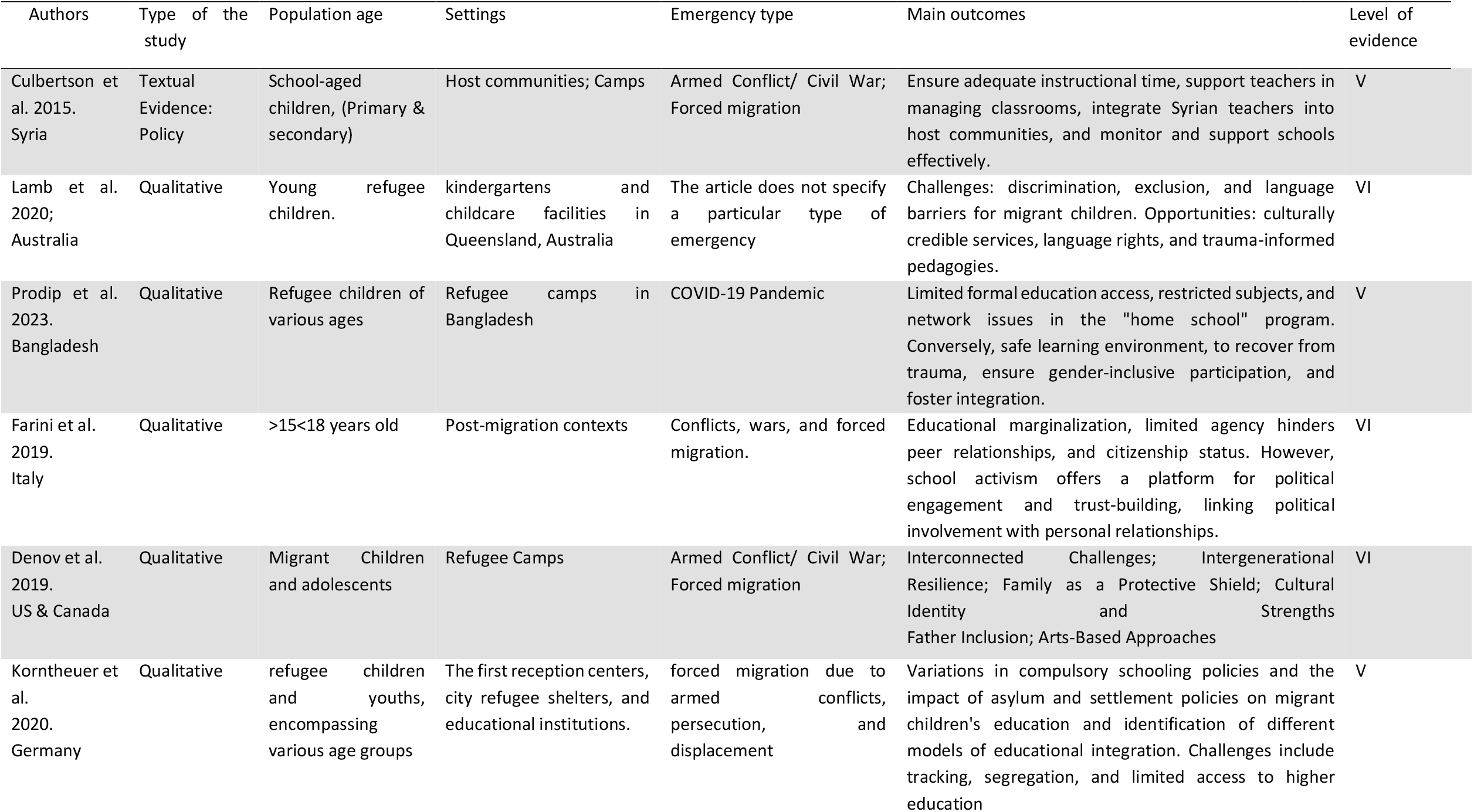
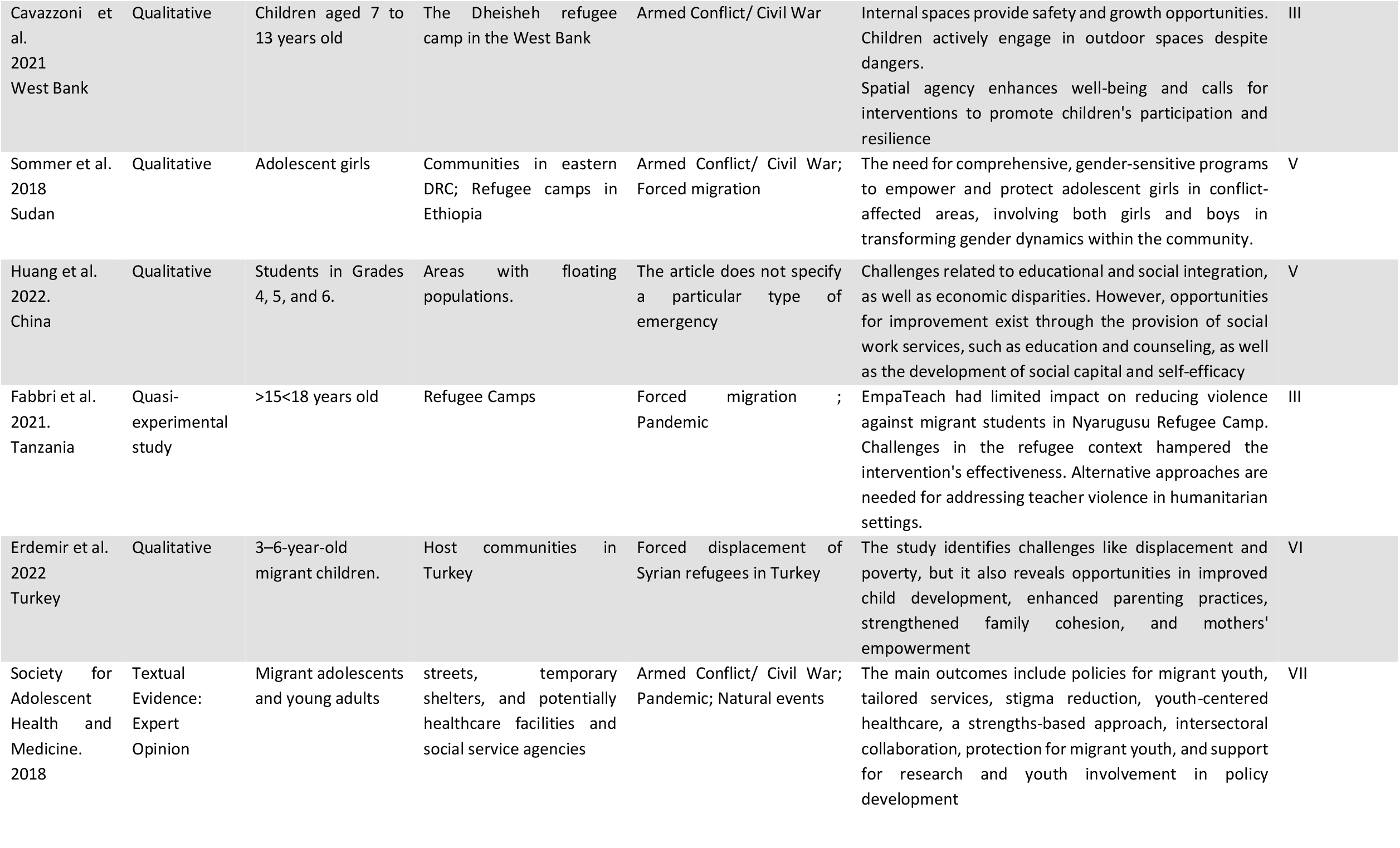
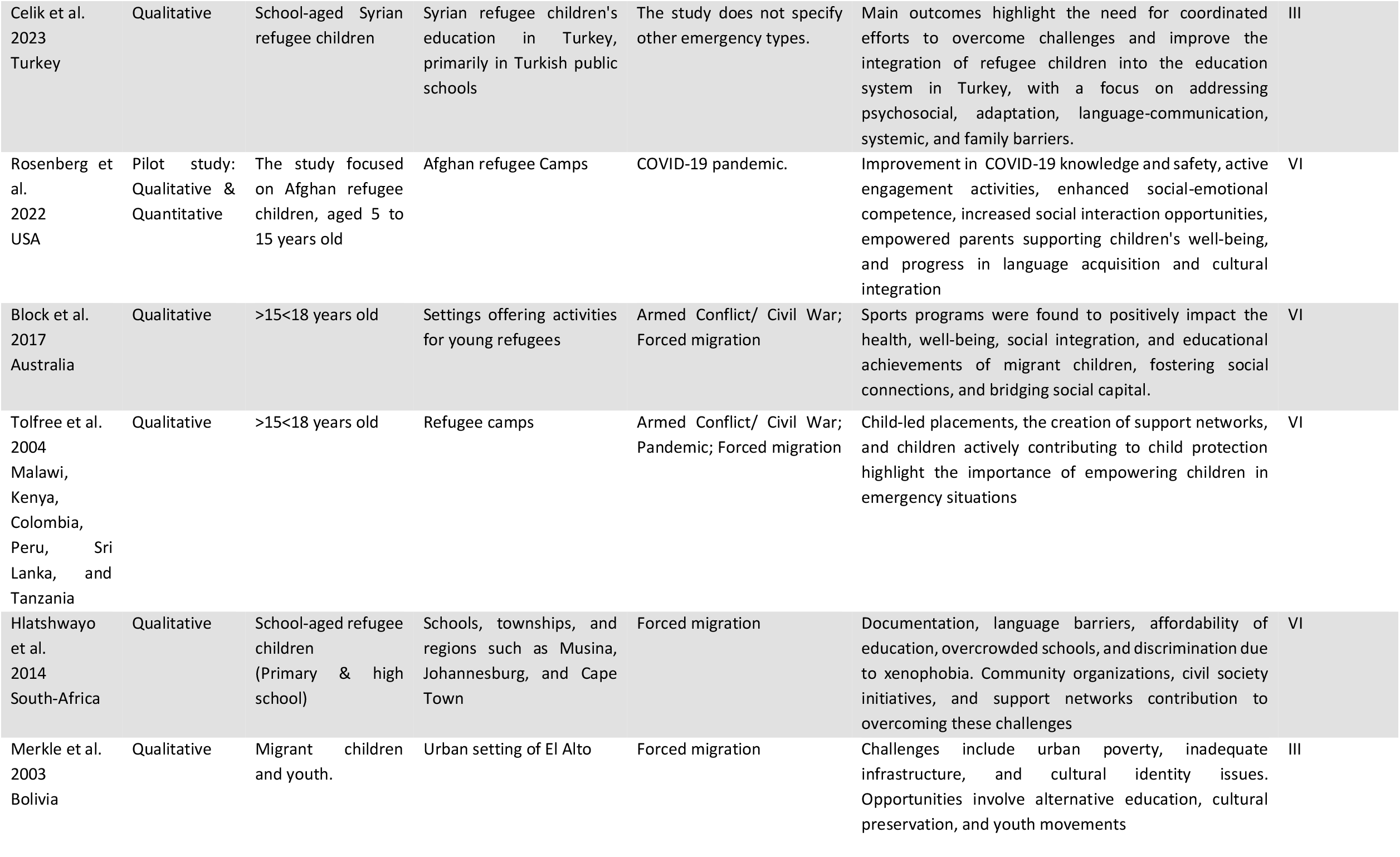
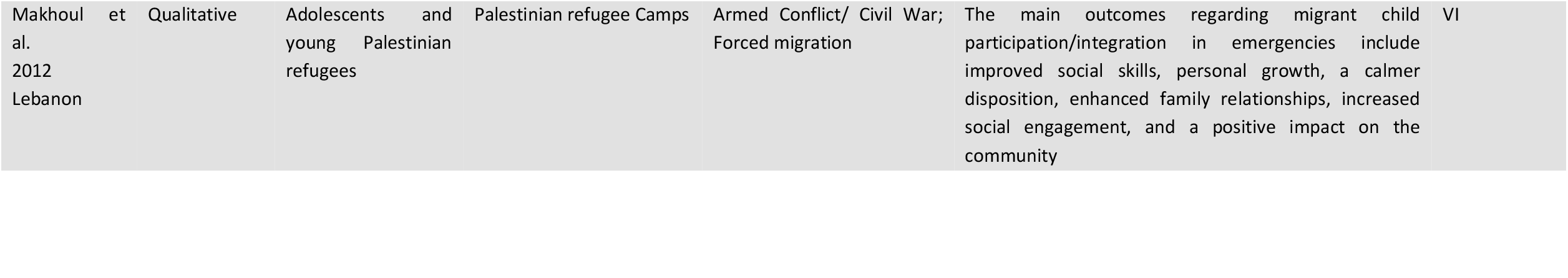
comprehensive summary of study characteristics.

