## Supplementary material for "Migrant children participation during emergencies: A Scoping review of global challenges and opportunities": Search Strategy

### The Role of Child Participation in Emergency Response for Internally Displaced Persons (IDP) Or Migrant Children in emergency and recovery response efforts: A Scoping Review Protocol of The Opportunities and Challenges

#### Objective:

The objective of this scoping review is to map the existing literature and identify the opportunities and challenges associated with child participation during emergency and response recovery effort, with a focus on internally displaced persons (IDP) or migrant children.

#### Review Question (s)

What are the opportunities and challenges associated with child participation during emergency and response recovery efforts, with a focus on internally displaced persons (IDP) or migrant children?

#### Definition of key concepts

##### The term child participation

The concept "**Child participation**" for this scoping review is defined by The State of the World's Children 2003 reports on child participation — the 'right' of all children to have their opinions considered when decisions are being made that affect them. UNICEF, 2003

This concept for this scoping review is going to be breakdown into the following terms to develop the search strategy:

- The term "**child**" for this scoping review is defined as:

1. **Child:**
   A person to 12 years of age. An individual 2 to 5 years old is CHILD, PRESCHOOL . PubMed
2. **Adolescent:**
   A person 13 to 18 years of age. PubMed

**Key Terms**: Children, Adolescents, Adolescence, Teens, Teen, Teenagers, Teenager, Youth, Youths, Female, Adolescent, Female, Female Adolescent, Female Adolescents, Adolescents, Male, Adolescent, Male, Male Adolescent, Male Adolescents

- The term "**Participation**" for this scoping review is defined based on the following key concepts :

1. **Social participation**
   Involvement in community activities or programs. PubMed, 2011
2. **Patient participation**
   Patient involvement in the decision-making process in matters pertaining to health. PubMed, 1978
3. **Community participation**
   Involvement of members of the community in the affairs of that community. PubMed, 1974
4. **Stakeholder participation**
   A process between an entity and those groups or individuals potentially or impacted by the actions of that entity over a range of activities and approaches. PubMed, 2018
5. **Empowerment:**
   Process of increasing the capacity of individuals or groups to make choices and to transform those choices into desired actions as deigned by the individuals or groups. PubMed, 2020
6. **Decision Making:**
   The process of making a selective intellectual judgment when presented with several complex alternatives consisting of several variables, and usually defining a course of action or an idea. PubMed

This concept for this scoping review is going to be breakdown into the following terms to develop the search strategy:

- The term "**Emergencies**" for this scoping review is defined as:

**Disasters**

Calamities producing great damage, loss of life, and distress. They include results of natural phenomena and man-made phenomena. Normal conditions of existence are disrupted, and the level of impact exceeds the capacity of the hazard-affected community. PubMed

**Armed Conflicts**

Any differences arising between two nations or groups and leading to the intervention of armed forces. PubMed

**Epidemics**
Sudden outbreaks of a disease in a country or region not previously recognized in that area, or a rapid increase in the number of new cases of a previous existing endemic disease. Epidemics can also refer to outbreaks of disease in animal or plant populations. PubMed
**Pandemics**
Epidemics of infectious disease that have spread to many countries, often more than one continent, and usually affecting a large number of people. PubMed

**Natural disaster**
Disasters linked to natural hazards including widespread fires, floods, storms, earthquakes, and drought. These events may result in significant damage and loss of lives. PubMed, 2019

Keywords: Emergency; disaster; earthquake; natural; Natural Disaster; Disaster, Natural; Seismic

- The term "**Emergency response**" for this scoping review is defined as:

1. **Disaster planning**
   Procedures outlined for the care of casualties and the maintenance of services in disasters. PubMed, 1978
2. **Relief work**
   Assistance, such as money, food, or shelter, given to the needy, aged, or victims of disaster. It is usually granted on a temporary basis. (From The American Heritage Dictionary, 2d college ed). PubMed, 1991
3. **Rescue Work**
   Activities devoted to freeing persons or animals from danger to life or well-being in accidents, fires, bombings, floods, earthquakes, other disasters, and life-threatening conditions. While usually performed by team efforts, rescue work is not restricted to organized services. PubMed, 1995

Key words: Planning, Disaster; Disaster Relief, Planning; Disaster Relief Plannings; Planning, Disaster Relief; Plannings, Disaster Relief; Relief Planning, Disaster; Relief Plannings, Disaster, Work, Rescue, Relief Works; Work, Relief; Works, Relief; Humanitarian Assistance; Assistance, Humanitarian; Assistances, Humanitarian; Humanitarian Assistances

#### The term “migrants and internally displaced persons (IDPs)” for this scoping review are defined as:

1. **Transients and Migrants:**
   People who frequently change their place of residence. PubMed

2. **Refugees**
   Persons fleeing to a place of safety, especially those who flee to a foreign country or power to escape danger, persecution, or economic distress in their own country or habitual residence. PubMed

3. **Disaster victims**
   Persons adversely effected by disasters, occurrences that result in property damage, deaths, and/or injuries to a community. PubMed, 2019

Keywords: Disaster Victim; “Victim, Disaster”, “Victims, Disaster” , Refugee; Political Asylum Seekers; Asylum Seeker, Political; Asylum Seekers, Political; Political Asylum Seeker; Seekers, Political Asylum; Political Refugees; Political; Refugee; Refugee, Political; Refugees, Political; Asylum Seekers; Asylum Seeker; Seeker, Asylum; Seekers, Asylum; Displaced Persons; Displaced Person; Person, Displaced; Persons, Displaced; Internally Displaced Persons; Displaced Person,; Internally; Displaced Persons, Internally; Internally Displaced Person; Migrants and Transients; Transients; Transient; Non-migrants; Non-migrant; Squatters; Squatter; Migrant Workers; Migrant Worker; Worker, Migrant; Workers, Migrant Migrants; Migrant; Nomads; Nomad

#### PCC Framework

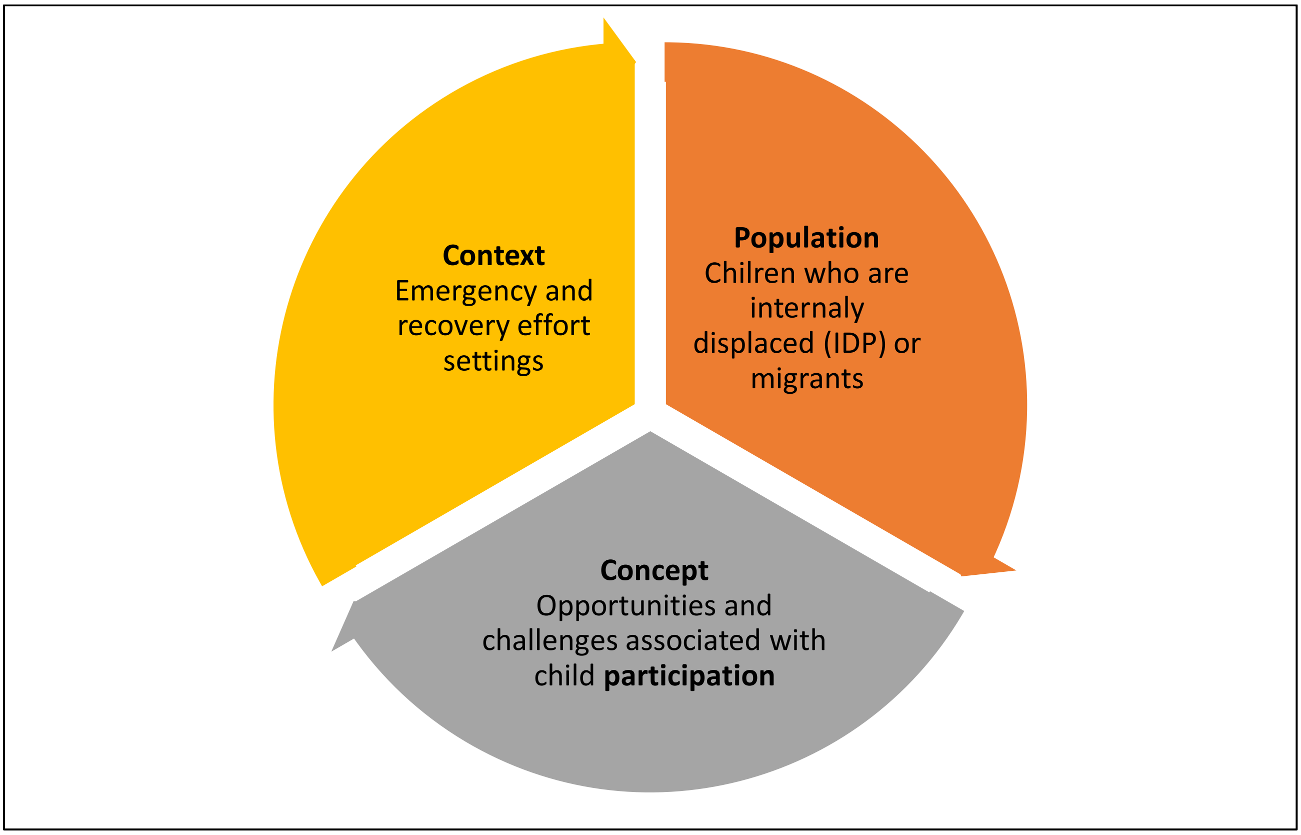


**This review is going to focus on the opportunities & challenges associated with the participation of migrants / IDP children in emergency and response effort settings**

#### Syntax PubMed [Mesh] [tiab]

| Search | Query | Results |
|---|---|---|
| #1 | **Concept 1: Child**<br>Child [MeSH Terms] OR adolescent [MeSH Terms] OR children [tiab] OR adolescen* [tiab] OR teen* [tiab] OR youth* [tiab] OR female [tiab] OR “female adolescen*” [tiab] OR “male adolescen*” [tiab] OR male [tiab] | **5,115,151** |
| #2 | **Concept 2: Participation**<br>“Social participation” [MeSH Terms] OR “patient participation” [MeSH Terms] OR “community participation” [MeSH Terms] OR “stakeholder participation” [MeSH Terms] OR empowerment [MeSH Terms] OR “decision making” [MeSH Terms] | 227,837 |
| #3 | **Concept 3: Post-Earthquake**<br>Disaster [MeSH Terms] OR Natural disaster [MeSH Terms] OR Earthquake [MeSH Terms] OR Emergencies [MeSH Terms] OR Emergenc* [tiab] OR disaster* [tiab] OR earthquake* [tiab] OR natural [tiab] OR seismic [tiab] | 1,285,727 |
| #4 | **Concept 4: Emergency Response**<br>“Disaster planning” [MeSH Terms] OR “relief work*” [MeSH Terms] OR “Rescue Work*” [MeSH Terms] OR Plan* [tiab] OR Disaster* [tiab] OR “disaster relief” [tiab] OR “disaster relief planning” [tiab] OR “planning disaster relief” [tiab] OR Rescue [tiab] OR “humanitarian assistance” [tiab] OR humanitarian [tiab] OR assistance [tiab] | 1,755,876 |
| #5 | **Concept 5: Migrants and IDPs**<br>“Transients and migrants” [MeSH Terms] OR Refugees [MeSH Terms] OR “Disaster victims” [MeSH Terms] OR “disaster victim*” [tiab] OR refugee [tiab] OR asylum [tiab] OR “asylum seeker*” [tiab] OR political [tiab] OR seeker [tiab] OR displaced [tiab] OR “displaced person” [tiab] OR “internally displaced person” [tiab] OR migrant* [tiab] OR transient [tiab] OR non-migrant [tiab] OR squatter* [tiab] OR “migrant worker*” [tiab] OR nomad* [tiab] | 464,488 |
| Result : **10 Articles** | | |

#### Syntax Scopus:

| Search | Query | Results |
|---|---|---|
| #1 | **Concept 1: Child**<br>TITLE-ABS (Child OR adolescent OR children OR adolescen* OR teen* OR youth* OR female OR "female adolescen*" OR "male adolescen*" OR male) | **5,171,095** |
| #2 | **Concept 2: Participation**<br>TITLE-ABS ("Social participation" OR "patient participation" OR "community participation" OR "stakeholder participation" OR empowerment OR "decision making") | **681,379** |
| #3 | **Concept 3: Post-Earthquake**<br>TITLE-ABS ( Disaster OR Natural disaster OR Earthquake OR Emergencies OR Emergenc* OR disaster* OR earthquake* OR natural OR seismic ) | **2,447,476** |
| #4 | **Concept 4 : Emergency Response**<br>TITLE-ABS ( "Disaster planning" OR "relief work*" OR "Rescue Work*" OR Plan* OR Disaster* OR "disaster relief" OR "disaster relief planning" OR "planning disaster relief" OR Rescue OR "humanitarian assistance" OR humanitarian OR assistance) | **6,391,734** |
| #5 | **Concept 5: Migrants and IDPs**<br>TITLE-ABS ("Transients and migrants" OR Refugees OR "Disaster victims" OR "disaster victim*" OR refugee OR asylum OR "asylum seeker*" OR political OR seeker OR displaced OR "displaced person" OR "internally displaced person" OR migrant* OR transient OR non-migrant OR squatter* OR "migrant worker*" OR nomad* ) | **1,818,360** |
| **Limited to:**<br>PUBYEAR > 2002 AND PUBYEAR < 2024 AND ( LIMIT TO ( LANGUAGE , "English" ) ) | | |
| Result :<br>**40 Articles** | | |

#### Syntax: Google Scholar

| Search | Query | Results |
|---|---|---|
| #1 | **Allintitle**: child participation AND Children OR Adolescent AND Migrant OR Refugee OR IDP AND Emergency response AND Turkey AND Post Earthquake | 1050 |
| **Limited to:** PUBYEAR > 1999 AND PUBYEAR < 2024 AND ( LIMIT-TO ( LANGUAGE , "English" ) ) | | |
| Result : **1050 Articles** | | |

#### Inclusion and exclusion criteria:

| Inclusion | Exclusion |
|---|---|
| Target population :<br>• Migrant Children<br>• Refugee Children<br>• Adolescents<br>• IDP | Population<br>• Adults<br>• Non-migrants |
| Concept of Study<br>• Participation in emergency | Concept of Study<br>• Any other |
| Context<br>• Emergency response<br>• Post- earthquake<br>• Turkey | Context<br>• Times of peace |
| Type of studies<br>• Original studies Qualitative<br>• Quantitative<br>• Mixed Methods<br>• Field reports | Type of studies<br>• Perspective<br>• Reviews<br>• Letter to editor<br>• Book reviews<br>• Case reports<br>• Policy guidelines reports |
| Region<br>• All | Region<br>• N/A |
| Language<br>• English | Language<br>• All other languages |
| Timeframe<br>• Last 20 years – Since 2003 | Timeframe<br>• Before 2003 |
